# Demographic profile, risk stratification, clinical characteristics, and treatment outcome of patients with multiple myeloma: A five-year retrospective study at a tertiary hospital in Ethiopia

**DOI:** 10.1101/2023.10.01.23295350

**Authors:** Ephrem Haile, Adugna Tasew, Amha Gebremedhin, Abdulaziz Sherif, Fissehatsion Tadesse

## Abstract

**Background:** Patients with multiple myeloma are being seen at an increasing frequency in different indigenous African population. However, local data regarding the demographic profile, clinical characteristics, risk stratification, and treatment outcome of these patients is lacking. This study was designed to fill this existing gap in our setup. Hence, it will aid in the revision of treatment guidelines based on local data on the efficacy of existing treatment regimens and risk stratification of patients with a newly diagnosed multiple myeloma.

**Methods:** A single centered Hospital-based retrospective Cohort study was conducted from January 2015 to December 2019. Eighty patients with newly diagnosed MM who received non-proteasome inhibitor-based therapy at TASH, Addis Ababa, Ethiopia were analyzed in the study.

**Results:** Out of the 80 patients in this cohort, 51(63.8%) of the patients were males (M: F ratio 1.76:1) and the median age at diagnosis was 52 years. The commonest complications identified were anemia (56.3%) and pathologic fracture (55%). The commonest comorbid conditions were; systemic hypertension (24%), CKD (6.3%), and diabetes (5%). The median Progression-Free Survival (PFS) and Overall Survival (OS) of patients were found to be 17.5 and 20 months respectively. This study also identified factors like advanced DS stage, presence of plasmacytoma, renal dysfunction, elevated serum LDH, high levels of serum protein, and monoclonal M-protein to have adverse implication on the OS and PFS of patients.

**Conclusion:** Multiple Myeloma is more common in the male population group and our patients are younger than the western population. Myeloma treatment regimens like CP and CPT are found to be less effective in our patients than in patients elsewhere. This is likely to be due to the advanced stage at presentation. In resource-limited setups, where determination of cytogenetic features of myeloma is difficult, different clinical and laboratory parameters can still serve as prognostic markers of treatment outcome & patient survival.

**Competing Interests:** The authors declare that they have no competing interests

## Background

With increasing life expectancy and the aging of the population, the prevalence and incidence of multiple myeloma and related plasma cell disorders are currently rising to become an important public health burden worldwide. (1) Myeloma accounts for approximately 1% of all malignant disorders and 10% of hematologic malignancies. In the western world, the age-standardized incidence is reported to be approximately 5 cases per 100,000 populations. (2) The Median age at diagnosis is reported to be 66 years with only 2% of patients being less than the age of 40 years at diagnosis. Men are affected by multiple myeloma more frequently than women (3) and individuals of African descent have twice the prevalence of MM as those of European descent. Though local national data on the magnitude of the disease is lacking, a study done by *Diana Oelofse et al,* in the indigenous African population suggested a progressively rising incidence and that MM accounts for around 13% of diagnosed cases of Hematologic malignancies. (4)

Multiple myeloma is a disease, which had been uniformly fatal within a few months of diagnosis. With the introduction of novel potential therapeutic agents and efficient stem cell transplant, significant progress has been made in the treatment of patients with myeloma. (5) Even though evolving treatment regimens over the years prolonged survival, they are unsatisfactory in giving a cure or a lasting treatment response. Therefore, the treatment of multiple myeloma is still continuously evolving.

IMWG consensus on risk stratification in multiple myeloma uses various prognostic markers in the risk stratification of patients with multiple myeloma. These include; the presences of specific cytogenetic abnormalities, the extent of the disease as evidenced by appropriate imaging techniques, the serum-free light-chain level, and the use of the International Staging System (ISS). **(6)** Robert A Kyle et al, also showed that complications and clinical presentations of myeloma including; severe anemia, infections, VTE, renal and extent of bone diseases can significantly compromise the quality of life of patients and could be important prognostic indicators determining treatment outcome and survival. (3) Hence, beside initiation of a selected appropriate treatment regimen in patients, follow-up, early identification and management of specific complications and organ dysfunctions associated with myeloma should not be overlooked.

With the introduction of novel agents and the availability of ASCT, survival rates of MM improved significantly. However, access to effective care is very limited in low-income countries, particularly in sub-Saharan Africa. In developing countries like Ethiopia where potent novel agents of myeloma like Proteasome inhibitors are just recently introduced and standard stem cell transplant for eligible patients is not available, the extent of treatment response and survival with the existing treatment regimens, mainly involving IMIDs, needs to be studied. Besides, in these resource-limited setups where internationally proven and accepted prognostic markers of multiple myeloma could not be applied into clinical practice, it would be reasonable to investigate and identify alternative clinical parameters determining treatment outcome and survival of patients.

Local data regarding the magnitude, epidemiology, survival, and treatment outcome of patients with the disorder is lacking. Therefore, this study aims to provide a single-center experience regarding the demographic & clinical characteristics, treatment outcome, and important clinical prognostic factors determining the survival of patients managed at TASH, Addis Ababa, Ethiopia. This study will aid in the revision of treatment regimens based on local data on the efficacy of treatment agents and risk stratification of patients with a newly diagnosed Myeloma.

## Methods

This single centered Hospital-based retrospective cohort study was conducted in the outpatient department of the Hematology unit in *Tikur Anbessa specialized referral and Teaching Hospital,* one of the largest teaching hospitals in Ethiopia. The study period for this retrospective study was from January 2015 to December 2019. The study aims to assess the patient demographics, clinical features, risk stratification, and determinants of treatment outcome in patients with a confirmed newly diagnosed multiple myeloma. All observed cases in the study period were included in this retrospective cohort study. 92 patients with confirmed multiple myeloma, according to the IMWG diagnostic criteria, and on follow-up at the outpatient hematology clinic of TASH in the specified study period were assessed for illegibility. 4 Patients with incomplete workup & laboratory data on diagnosis and follow-up and 3 patients for whom specific chemotherapy was not initiated or the period of follow up is less than three months were excluded from the study. As the primary objective of this study was to assess the treatment outcome of patients who received alkylator-based regimens used in resource-limited setups, 5 patients who were managed with Proteasome inhibitor-based combination chemotherapy were also excluded from the study. 80 patients with a newly diagnosed MM were finally included in the data analysis.

International staging system (ISS) of MM could not be used as determination of cytogenetic features and serum B_2_ microglobulin levels of patients is not readily available in resource limited setups like Ethiopia. Patients were accordingly staged based on the revised Durie-Salmon clinical staging criteria. Remission induction treatment options available for patients were the CP (Cyclophosphamide – Prednisolone), CPT (Cyclophosphamide – Prednisolone-Thalidomide) MPT (Melphalan - Prednisolone-Thalidomide) regimens. Serum and urinary M-protein immune-fixation & FLC ratio could not be determined; and hence IMWG response criteria was not applicable in our setup. We used the median OS and PFS of patients to assess treatment outcome in patients. None of the patients received stem cell transplant following remission induction treatment.

Clinical records and laboratory data available for all illegible patients on follow-up and treatment in the outpatient unit were reviewed. A structured data collection format was used to fill in data obtained from patients’ chart records. Some laboratory data were also obtained from electronic data recording of the hospital’s Laboratory. Data were then processed and analyzed using SPSS (V26) statistical software. We used descriptive statistics parameters to assess the association between the study variables. Kaplan-Meier estimator was used to analyze the OS and PFS of the patients using different variables. In all cases, a p-value of less than 0.05 was considered to depict a statistically significant association among study variables.

## Results

### Demographic characteristics of Study participants

Out of the 80 patients, 51 (63.8%) were Males and 29 (36.2%) were females. The Median age of patients at diagnosis was 52 years. The mean age at diagnosis was 54 years (range 25 – 82). The majority of the patients (62.5%) were between the age of 40 years and 65 years. Extremes of age, less than the age of 40 years and greater than the age of 65 years, were each encompassing 18.8% of the study participants. Patients were seen from all regions of the country except for the regions of Gambela and Afar. In this cohort, 35% of the patients were from Addis Ababa and 33.7% & 13.7% of the patients were from Oromia and Amhara regions respectively. The other 4 regions contribute to the remaining 19% of the patients. (Table 1)

### Clinical Characteristics of study participants

Patients in the study were managed with 3 major induction regimens; which include the CP (Cyclophosphamide – Prednisolone), CPT (Cyclophosphamide – Prednisolone-Thalidomide), MPT (Melphalan - Prednisolone-Thalidomide) regimens. Sixty percent of the patients were given the CP induction treatment regimen while 32.5% and 6.3% of patients were managed with CPT and MPT regimens respectively. Only a single patient was given a TP (Thalidomide-Prednisolone) combination Therapy. Apart from decisions from physicians in holding treatment for various treatment-related complications, 90% of the patients completed their induction treatment course with no discontinuation of therapy or follow-up.

One or more major comorbidities were identified in 36% of the patients. These include Systemic Hypertension (24%), CKD (6.3%), and Diabetes mellitus (5%). Only a single patient was found to have an underlying cardiac illness, IHD. Fifty-nine percent of patients had no known underlying comorbidity detected. (Figure1)

Evaluation for the disease stage at diagnosis based on the DS (Durie-Salmon) clinical staging criteria was also made. The majority (61.3%) of the patients had a stage IIIA disease followed by a stage IIA accounting for 23.8% of the cases. Male patients in the study, as compared to females, presented at advanced stages but the Mann Whitney test of medians didn’t show a level of statistical significance (P= 0.13). (Figure 2)

Renal failure in the staging process was seen in 15% of study participants at the presentation. Lytic lesions visualized on a plain x-ray were present in 93.8 % of diagnosed patients and only 5% of patients did not develop this in the follow-up period.

### Laboratory Features of Patients in the study

57.3% of the cases had anemia of different degrees of severity at diagnosis and an additional 5.2% developed the condition in the follow-up period. The mean hemoglobin level at diagnosis was 10.13 g/dl. Twenty-five and Thirty-one percent of patients had severe and moderate anemia respectively. Sixteen percent of the patients required blood transfusion for anemia while only two patients were given EPO therapy for persistent and refractory anemia affecting patient management. Anemia was absent in 37.5% of the patients. (Table 2)

Hypercalcemia was another common laboratory parameter identified in the study. 37.6 % of patients had a serum calcium level, adjusted for the degree of hypoalbuminemia, above the upper normal level with a mean value of 10.2 ± 2.4 mg/dl. Only a single patient developed new-onset hypercalcemia after diagnosis in the study period.

Renal failure was documented in 15% of the study participants at presentation and even more proportion (18.8%) of patients had a creatinine value above the laboratory UNL. Five patients with renal dysfunction required a RRT and none of these patients had a recovery in their kidney function. 73.8% of the cases had a normal renal function determination throughout the follow-up period.

From serum protein electrophoresis results, the mean value for monoclonal ‘M’ and also total protein were 4.2 g/dl and 9.6g/dl respectively. Fifty-six percent of the patients had hypoalbuminemia and the mean serum albumin level was 3.1± 0.7g/dl. The mean bone marrow plasma cell percentage was 27±20.5%. LDH level was elevated in 76.3 % of patients with a mean value of 373.4 ± 237.6 U/L at the time of disease diagnosis.

### Treatment and disease-related Complications

Peripheral neuropathy developed in 16.4% of the patients on follow up and patients who received thalidomide containing regimen had a statistically significant more risk (P= 0.04) and Odds ratio of 3.4 (95% C.I, (1.01-11.2)) for the development of peripheral neuropathy during follow up.

A Significant number of patients had a major neurologic complication including neurologic weakness and bladder dysfunction. 31.3% of study participants developed one of these complications and Para paresis was the most common encompassing 76.2% of the neurologic complications seen in patients. The development of neurologic weakness had a statistically significant correlation with the presence of pathologic fracture (P= 0.001) and plasmacytoma (P=0.02). 44(55%) and 25(31.3%) patients had a documented pathologic fracture and an osseous or extra medullary plasmacytoma respectively.

Venous thrombosis was seen only in 6.3% of the study participants. Eighty percent of the cases with venous thrombosis were females. Thrombotic episodes occurred in both Thalidomide and Non-Thalidomide containing regimens. The number of thrombotic events was few for determination of its possible association with the type of regimen and the use of a VTE prophylaxis.

Infections, all of which were cases of pneumonia, were identified in 41.3% of patients on follow-up. A Statistically Significant variation was not detected in the risk of development of pneumonia among the different treatment regimens. Only a few (4) patients received antibiotic prophylaxis for a correlation to be made with its benefit of infection prevention.

It was difficult to determine common causes of death in patients managed for myeloma in this particular study as most of the patients died outside of the hospital and there was no tracking system for others who are lost to follow up after completion of their induction treatment. Out of the 20 patients for whom the immediate cause of death is documented; disease relapse or refractory disease, renal failure, and severe pneumonia with related complications made up the 3 most common in Hospital causes of mortality; accounting for 35%, 30%, and 25% of the cases respectively

### Treatment Outcome and determinant Factors

The outcome of treatment for myeloma, in this particular study, was assessed through the overall Survival (OS) and Progression-free survival (PFS) of patients. The median OS of patients was 20.0 months. Twenty five percent of the patients had a survival period not exceeding 9 months while 25% survived for more than 45 months. The 02-year and 03-year OS of patients was 50% and 28.6% respectively. The median PFS or period of an SD was 17.5 months. The 02-year and 03- year PFS of patients was 36.3% and 23.8% respectively. (Figure 3)

Different patient, disease, and treatment-related characteristics were assessed as a determinant factor for the treatment outcome of patients. Demographic features like gender and age were assessed but survival did not tend to differ, with statistical significance, by gender or across the different age groups. Pearson’s method of correlation showed a negative correlation between age and survival but it was not statistically significant. The negative correlation between age and Survival is strong in the first 01 years of follow-up; and once patients survived past the first year, the contribution of age in survival lessens out.

Treatment-related factors like VTE prophylaxis usage, a requirement for blood component transfusion, use of prophylactic antibiotics with chemotherapy were found to have no association with OS or PFS of patients.

D-S disease stage at presentation was significantly skewed as most patients presented with stage III disease. As no patient was diagnosed with a stage I disease, patients were categorized into a stage II and a stage III disease at diagnosis. The non-parametric Spearman correlation coefficient between disease stage and survival showed a negative correlation. As compared to patients with a stage III disease, patients diagnosed with a stage II disease had a statistically significant better 01 year OS (P=0.03). The 01 year OS of patients with a stage II disease was 76.2% and this number significantly declines to 55.9% in patients with a stage III disease. This association was not found to be significant once patients survived past the first year of therapy. Kaplan Meier survival analysis didn’t show a statistically significant association between disease stage at diagnosis and PFS throughout the follow-up period. (Figure 4)

Pathologic fracture at presentation or encountered in the follow-up period, was more common in males but the association was not strong enough to reach a level of statistical significance (P= 0.24). Though bisphosphonate therapy didn’t have a survival benefit demonstrated in this study, its ability in preventing a new-onset lytic lesion or pathologic fracture was significant (P=0.01).

Total serum protein from the SPEP was found to have a statistically significant correlation with both OS and PFS with P values of 0.008, 95%C.I (-9.29, -1.56) and p=0.006, 95%C.I (-4.71, - 0.83) respectively. A 1g/dl rise in serum total protein was associated with a 5.4 and 2.8 months decline in OS and PFS respectively. A similar degree of significant association was also shown between serum M protein levels and survival. (Table 3). A 1 g/dl rise in serum monoclonal M protein resulted in a 2.7 and 2.9 months decline in PFS and OS of patients respectively.

Treatment and disease-related complications were also assessed for their prognostic implications. Venous thrombosis, development of pathologic fracture & neurologic weakness, neutropenia, and also the development of pneumonia failed to demonstrate a statistically significant association with survival. Survival outcomes also tend not to differ in individuals with documented major comorbidities (hypertension, diabetes), assessed in the study, as compared to those with no such conditions.

The presence of Plasmacytoma in our patients with MM had a statistically significant negative association with OS (P= 0.046, 95% C.I (-21.47,-.211) of patients. The interaction between the presence of an extra medullary disease and survival was not found to be significant in the first 02 years of follow-up. However, in the consequent periods of follow-up, the OS of patients was significantly influenced by the presence of extra medullary disease at diagnosis. The three-year OS of patients with no coexisting plasmacytoma was 31% and this number significantly declines to 12% in patients with a plasmacytoma. (Figure 6)

Serum LDH level was the other disease activity marker shown to have a prognostic impact on survival. Seventy-six percent of the study participants had an elevated serum LDH value at presentation. A statistically significant level of negative correlation was seen between serum LDH level at diagnosis and OS of patients in the study (P= 0.02, 95%CI (0.007-0.078). The linear regression association between the variables shows for every 100U/L rise in serum LDH, the OS of patients dropped by 4.3 months. A similar significant association was not demonstrated with the serum LDH level at diagnosis and PFS of patients. (Table 3)

18.8% of patients had a deranged renal function test at presentation. Based on their baseline serum creatinine patients were divided into 3 groups; those with serum Cr within the laboratory normal range, UNL – 2mg/dl, and above 2mg/dl. Kaplan Meier survival analysis showed a statistically significant negative correlation between derangement in renal function and both OS and PFS of patients. Patients with normal serum creatinine at presentation had a 01 year and 02 years OS of 62.3% and 57.4% respectively. This figure drops significantly in patients with serum creatinine in the range of 1.2 – 2mg/dl to a 28.6% 02 years OS (P=0.01). Patients with serum creatinine more than 2mg/dl at presentation also had a significant drop in 02 years OS to 25%. (Figure 5)

## Discussion

Out of the 80 patients identified in this five-year retrospective cohort, 63.8% of them were males (M: F ratio 1.76:1), which is similar to the strong male preponderance of the disease reported from different studies. (3) Patients in our study were found to be younger (Median: 52 years) than the western population. The number of patients diagnosed before the age of 40 had a significantly higher proportion (18.7%) when compared to retrospective review of patients with newly diagnosed myeloma at the Mayo clinic by *Robert A Kyle et al*. (3)

The most common complication identified in the study was anemia (56.3%) & pathologic fracture (55%). The most common comorbid conditions identified were systemic hypertension (24%), CKD (6.3%), and Diabetes mellitus (5%). These conditions are also mentioned in studies by *Kleber M et al* and *Xue Song et al* as the most prevalent disease related complications & comorbidities. (7,8) Conforming to reports from these studies, results from our study also showed that comorbidities like hypertension and diabetes are not significantly related to survival during the study period. (8)

The prevalence of disease-related complications like pneumonia, renal, and bone disease in our patients was significantly higher than other similar studies in the United States and Europe. (7, 8) More than half of the patients (56.7%) in this study had anemia. This proportion is similar to reports from a study conducted by Nwabuko OC et al, in another similar African oncologic treatment center in Nigeria. (9)

Despite the introduction of Proteasome inhibitor-based regimens as the first-line treatment for newly diagnosed patients with myeloma, this study primarily intended to evaluate the efficacy of earlier regimens (CP, CPT and MPT) which are still used in the country. With the high treatment-related cost of newer agents and lack of alternative regimens in treatment failure, alkylators like Cyclophosphamide, are still widely used in resource-limited regions of the world.

The median PFS and OS of patients in this study were 17.5 and 20 months respectively. This is shorter than a report from a similar Sub Saharan Arican study in Niger Delta, Nigeria by Nwabuko OC et al, where the overall mean survival interval was 45 months for patients treated mainly with MP and MPT treatment regimens. In another similar myeloma trial study by Gareth J. Morgan et al, where patients received CTD regimen, the median OS of patients was 33 months. (9, 10)

This study showed a statistically significant negative correlation of survival with both renal dysfunction and disease stage at presentation. Patients with a D-S stage II disease, similar to other studies(11), were found to have better 01-year survival (76.2%) as compared to a D-S **s**tage III disease (55.9%). Patients with renal dysfunction at presentation had a significant drop in 01- year and 02 years OS from 62.3% to 28.6% (P=0.004) and from 57.4% to 25% (P=0.001) respectively. *F.A Fasola et al,* also demonstrated this poor prognosis and survival associated with acute kidney injury in patients with myeloma, at the Hematology department of a tertiary hospital in South-Western Nigeria. (13)

Total serum protein and serum monoclonal M protein levels had a statistically significant correlation with both OS and PFS of patients. This is in contrary to reports from other prognostic studies in myeloma which didn’t find a statistically significant association between these markers and patients’ survival. (12) The presence of osseous or extra medullary plasmacytoma also contributed to a significant decline in the 03 years and 04 years OS of patients from 31% to 12% (P=0.05) and from 27.3% to 8% (P=0.03). This finding is in concordance with reports from a retrospective cohort of newly diagnosed patients with myeloma at the University of Arkansas, USA. In this study, *S.Z. Usmani et al,* were able to demonstrate that, regardless of therapy, extra medullary disease was associated with shorter progression-free and overall survival. (14)

High levels of serum LDH at either diagnosis or following high dose chemotherapy was found, in some studies, to be an important factor associated with a shortened period of survival and correlate with variables like high serum B2 microglobulin level and renal dysfunction. (15,16) This study was also able to demonstrate that elevated serum LDH at diagnosis has a statistically significant association with reduced OS and PFS of patients with myeloma.

In this retrospective cohort, the negative correlation found in univariate analysis between survival and factors like age, hypercalcemia, marrow plasma cell percentage, and degree of anemia was not found to be statistically significant. This finding is consistent with results from the oncology department of the University of Pretoria (17) but in contrary to reports by *Michael E. Acquah1 et al,* a study conducted in a tertiary referral hospital in Ghana. (18) In this African retrospective study, unlike findings from this cohort; severe anemia, marrow plasmacytosis > 20% and hypercalcemia were shown to have significant impact on survival of patients with a newly diagnosed myeloma.

## Conclusion

Patients with MM in our setup were found to present in advanced stages of the disease. Physicians also need not overlook the incidence of the disease below the age of 40 years, as patients in our country are younger than the western population. Induction treatment regimens like CP and CPT were found to perform less in our patients than in other oncologic treatment centers elsewhere. Close follow-up, identification, and treatment of disease related complications is vital for a better treatment outcome. We identified factors like advanced DS stage, co-existence of Plasmacytoma, elevated serum LDH, high level of serum total protein, and monoclonal M protein to be important prognostic markers of patient survival. These factors can be utilized for risk stratification of patients in the absence of cytogenetic markers and other measures of disease burden like B2 microglobulin level.

To compare results with currently introduced Proteasome inhibitor-based therapeutic regimens, other similar follow-up studies are needed in our set up on patients who are and will be treated by these new regimens.

### List of Abbreviations

ASCT: Autologous Stem cell Transplant
CVD: Cardiovascular disease
CP: Cyclophosphamide – Prednisolone
CPT: Cyclophosphamide – Prednisolone-Thalidomide
D-S: Durie-Salmon
IMIDs: Immunomodulatory drugs
ISS: International Staging System
LDH: Lactate Dehydrogenase
MM: multiple myeloma
MPT: Melphalan – Prednisolone – Thalidomide
OS: Overall Survival
PFS: Progression-free Survival
TASH: Tikur Anbessa Specialized Hospital
TP: Thalidomide – Prednisolone
UNL: Upper normal limit
VTE: Venous Thromboembolism

## Declaration

## Ethical Approval and Consent to Participate

Ethical clearance was obtained from Addis Ababa University, College of Health Sciences Institutional Review Board. Consent was not taken from individual patients as it was a retrospective analysis. Only the data collectors recorded the patients’ data and the final combined data were accessible only to the principal investigator and authors. The information summarized is not discussed referring to the patient’s name.

## Consent for Publication

Not applicable

## Availability of Data Materials

The datasets used and/or analyzed during the current study are available from the corresponding author on reasonable request.

## Authors’ Contribution

The primary author was the principal investigator who did the conceptual framework, data collection, analysis and oversaw the research project. The co-authors were involved in data collection, analysis, interpretation, and reviewing several drafts of the research work.

## Data Availability

All data produced in the present study are available upon reasonable request to the authors

## Acknowledgments

We are very much grateful for all the help and support we got from the nurses and all the staff at the outpatient department of the unit of Hematology. We would also like to appreciate the staff at the hospital’s Patient Chart record room for the dedicated service we got during the data collection process. We are also very much thankful for all the support and understanding of our patients with whom we needed to communicate while conducting this study.

---

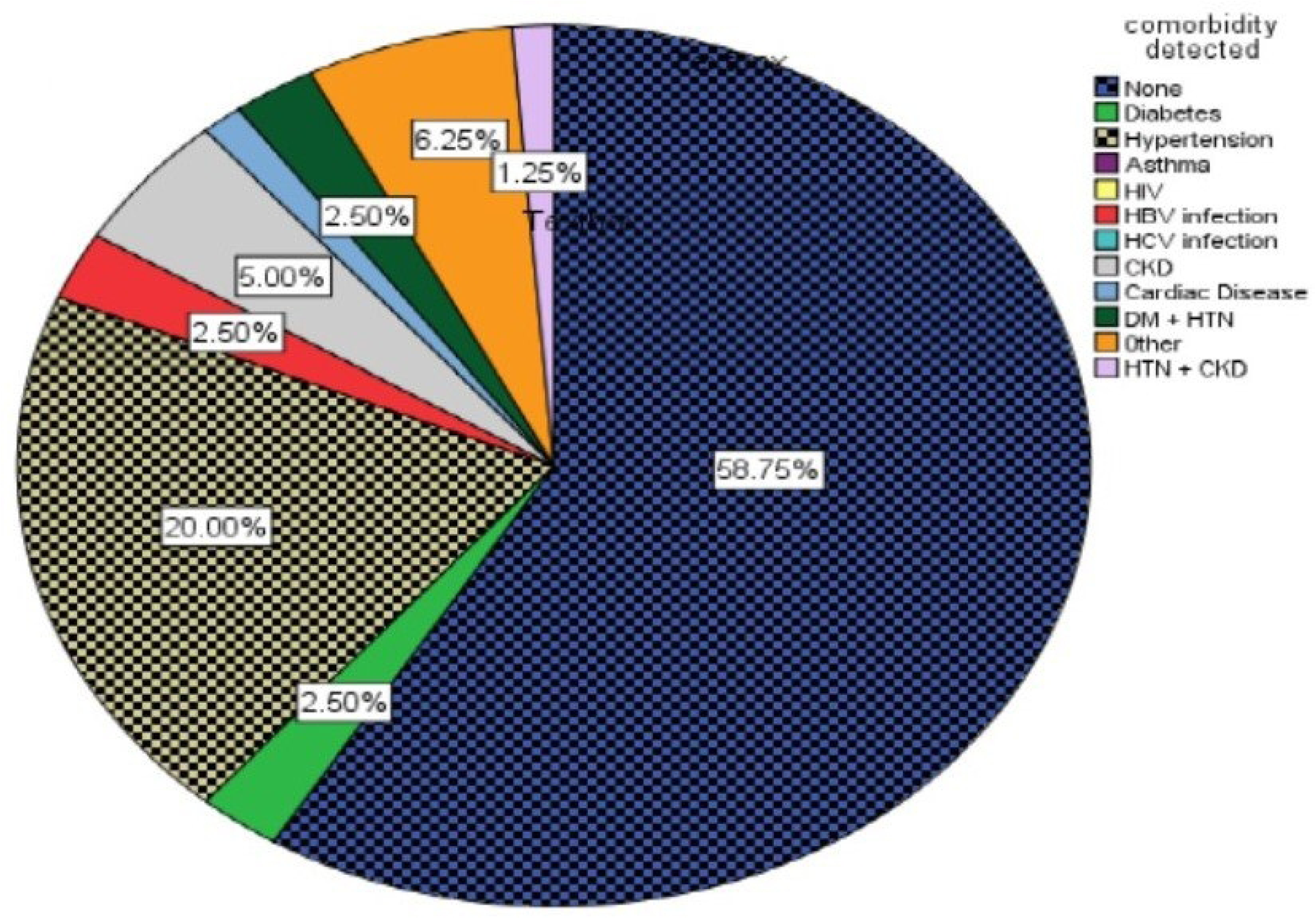

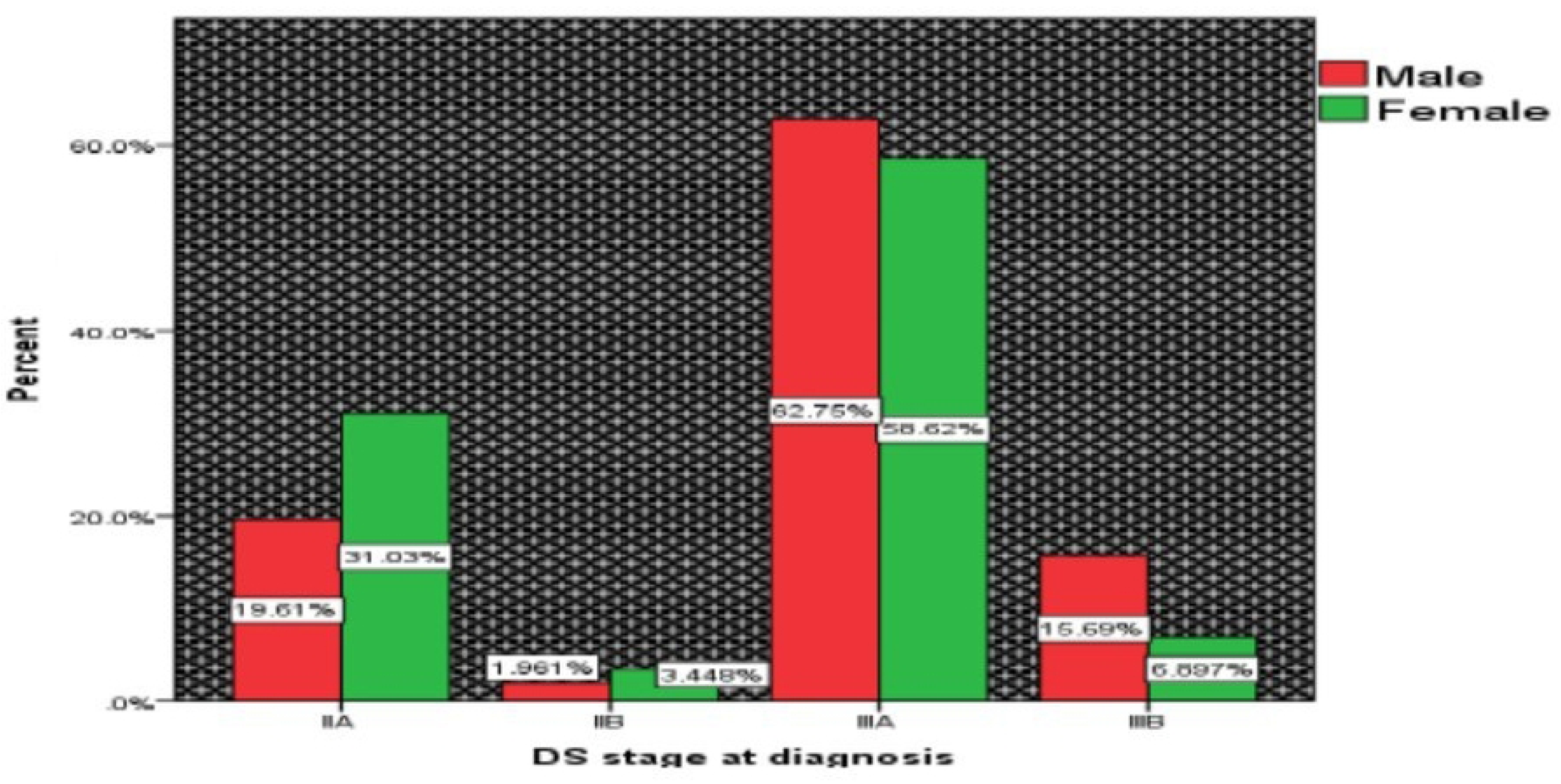

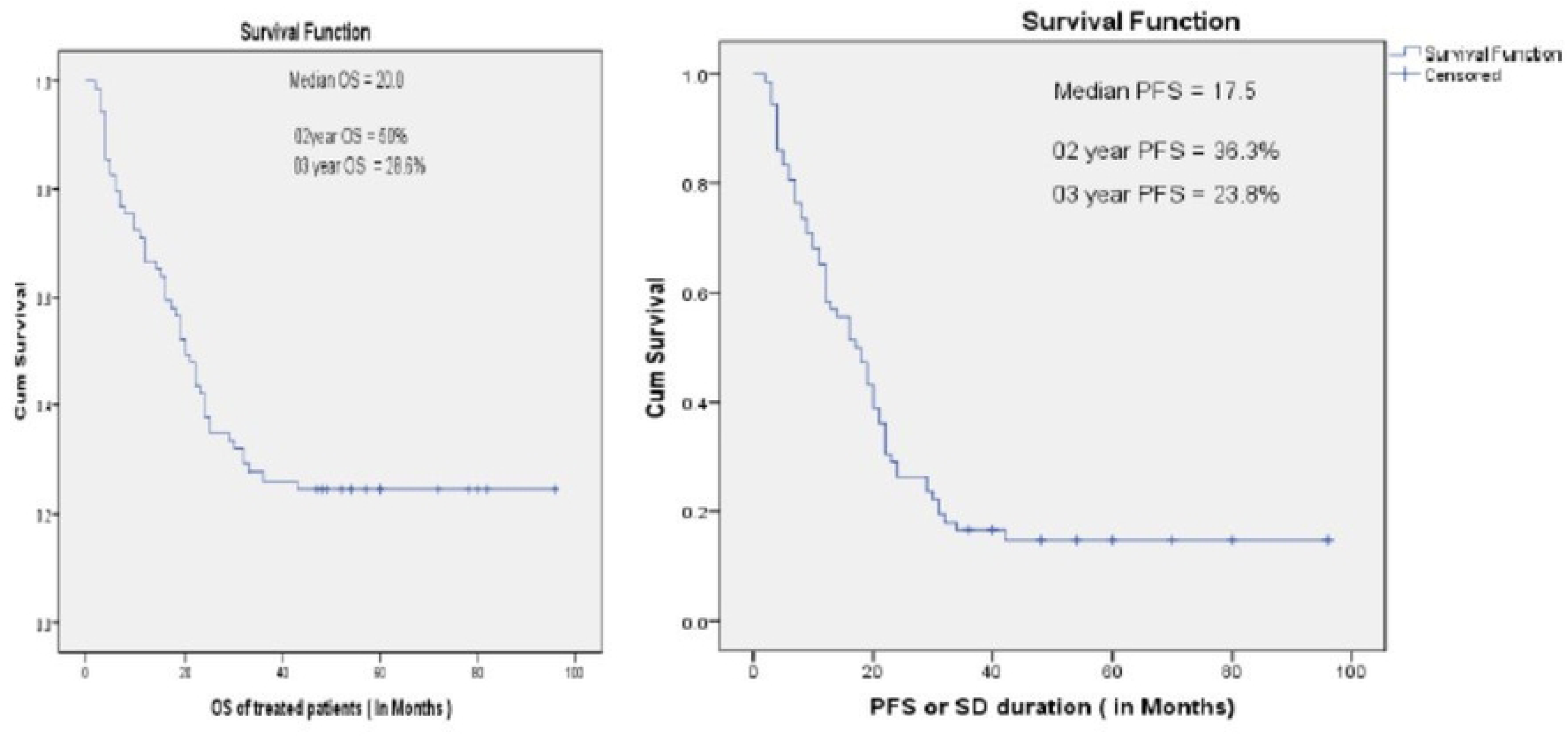

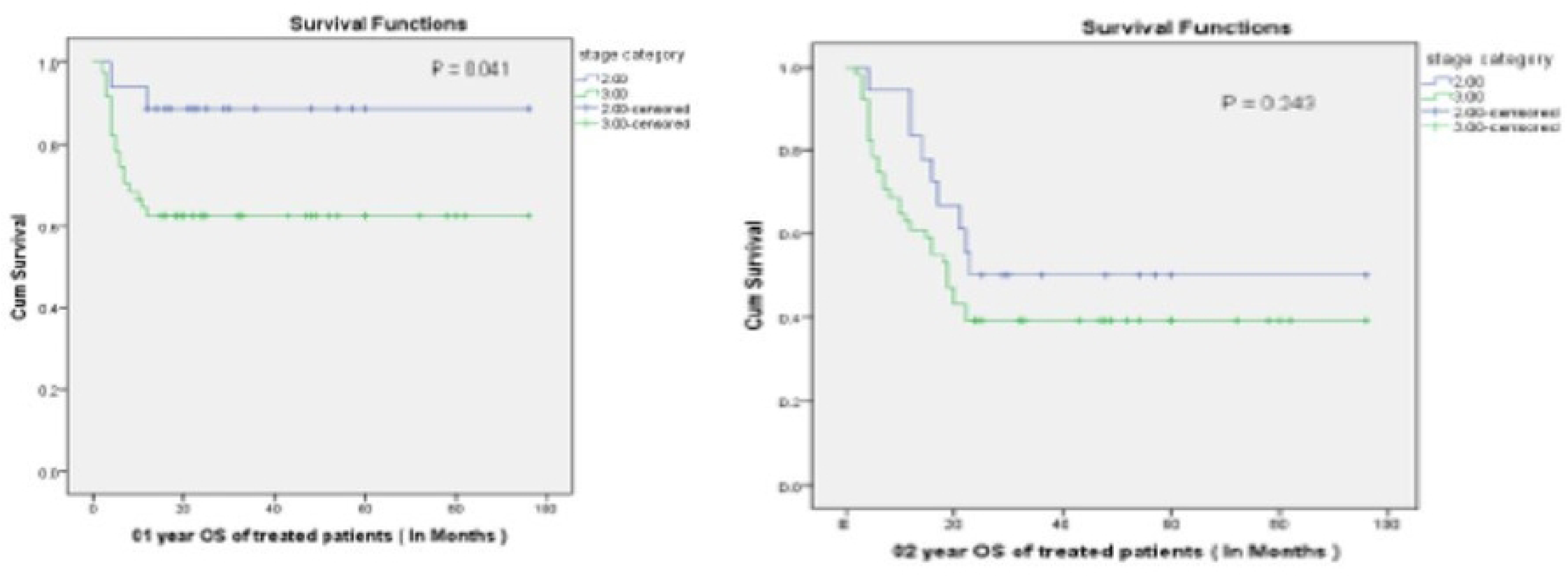

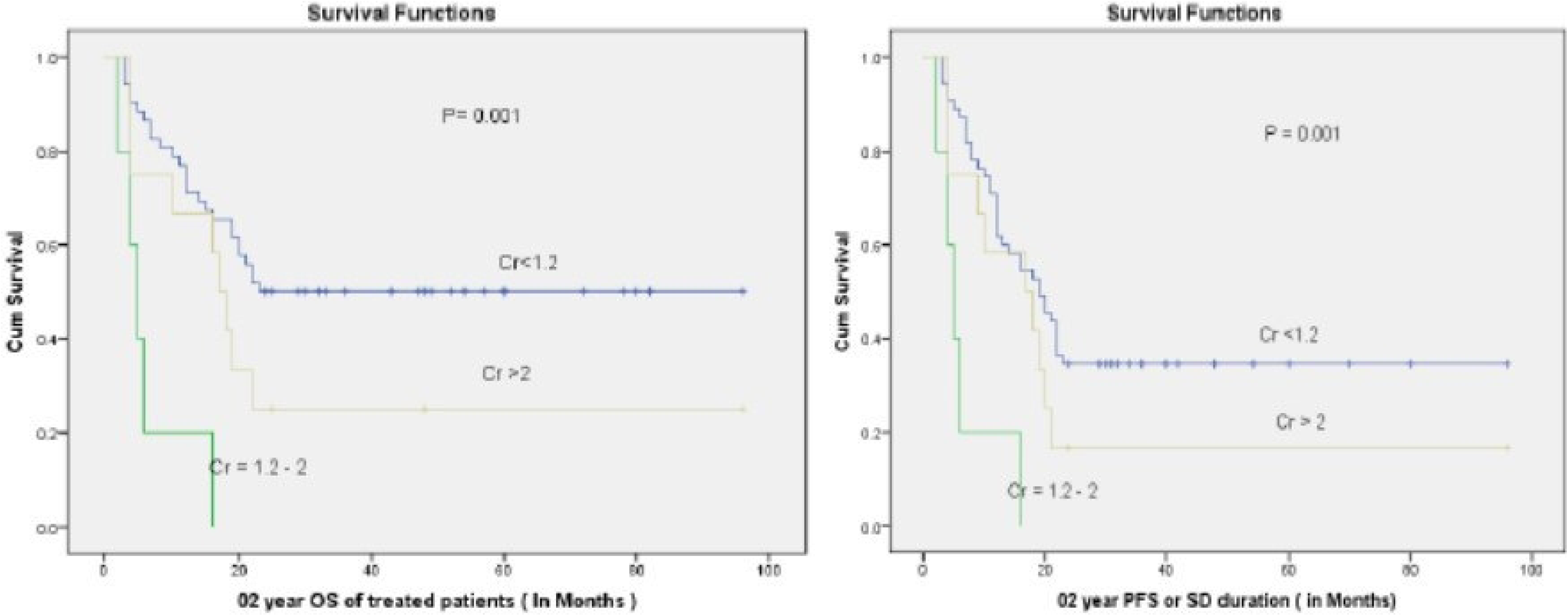

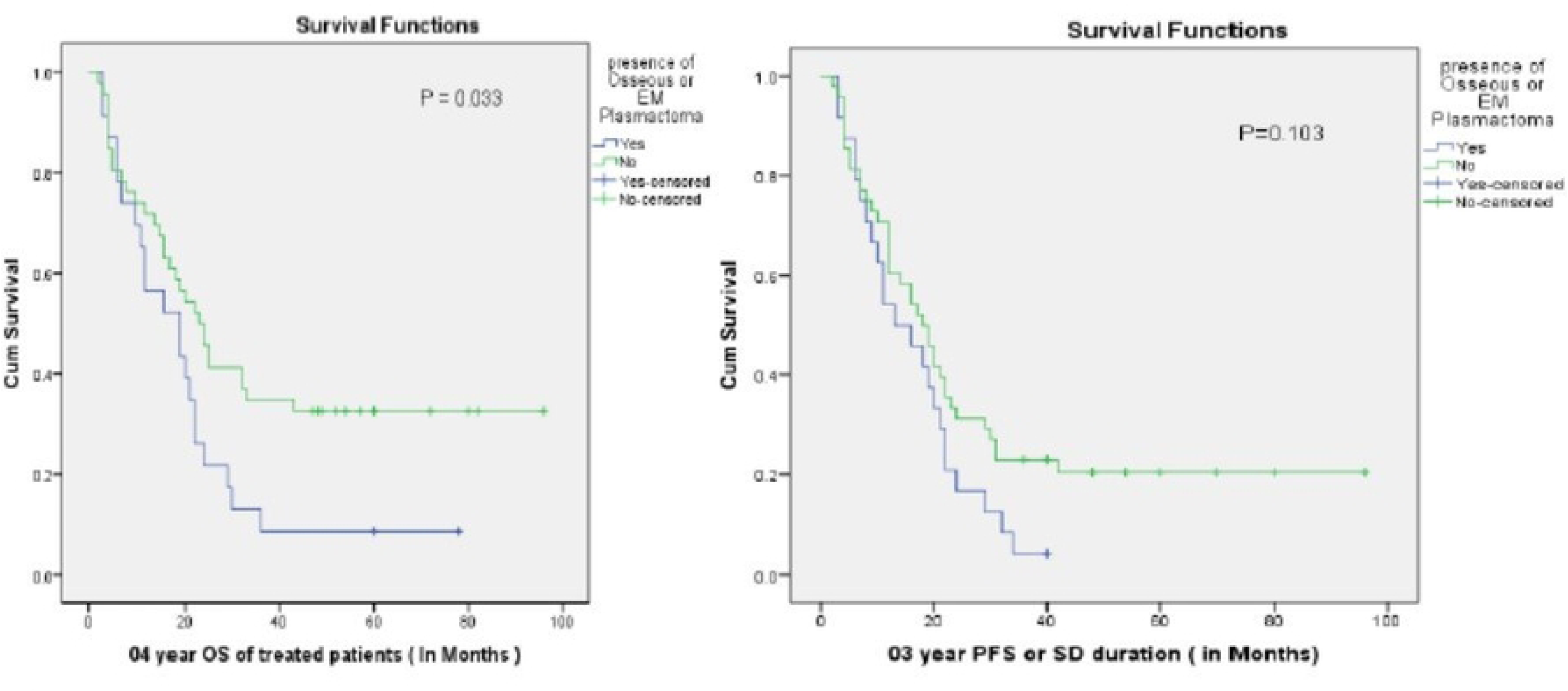

